# Deteriorated Covid19 control due to delayed lockdown resulting from strategic interactions between Governments and oppositions

**DOI:** 10.1101/2020.05.26.20112946

**Authors:** Alessio Carrozzo-Magli, Alberto d’Onofrio, Piero Manfredi

## Abstract

**Background:** In many European countries and the US, the burden of Covid-19 epidemic could be much lower if Governments had been able to learn from the China and Lombardy stories and to declare full lockdown without delays.

**Methods:** We use a simple game-theoretic framework for the strategic interaction between the Government, political oppositions and lobbies, combined with a Covid-19 transmission model, to analyse the role of political factors delaying the lockdown declaration, depending on the degrees of “responsibility” of political actors.

**Results:** The lockdown can always be declared immediately (i.e., without delay) as sustained transmission arises, only if the government feels fully “responsible” towards all citizens. If this is not the case, epidemic growth will eventually dominate the agents’ payoffs, so that sooner or later the lockdown will always be declared i.e., both the government and the opposition will be forced by the epidemic to switch towards a higher degree of responsibility, but with a delay. There is a further nontrivial situation where the lockdown can be declared without delay, occurring when the political opposition is at least as responsible as the Government. This however requires the solution of a coordination issue, which cannot be taken for granted. Eventually, a vicious circle emerges, where the delayed lockdown requires a much longer lockdown period to achieve adequate control results, thereby causing the explosion of economic losses and so calling for unlocking long before it should.

**Conclusions:** Lockdown delays have dramatically worsened the impact of the current Covid-19 wave in a number of countries. Citizens should be made cogently aware of this to claim maximal responsibility from political actors and economic lobbies to avoid that such stories repeat in the future when further threats, due to Covid-19 itself or other pathogens, will re-appear.

## 1. Introduction

After its explosion in Wuhan, in the Hubei province of China, documented since mid January 2020, the Covid-19 pandemic is ravaging the planet (WHO situation reports, 2020). Notwithstanding the fast dissemination efforts of WHO towards the world governments (Who Situation report 1, 20 Jan 2020), heterogeneous, often inadequate, interventions were implemented worldwide as first cases were locally notified. The first large epidemic in Europe occurred in Northern Italy, particularly in Lombardy region with its devastating toll, subsequently followed by Spain, France, the UK. In the afflicted Lombardy sites, the diagnostic systems, hospitals and ICUs have been largely overwhelmed, resulting in a massive mortality bulge (ISTAT, April 2020).

A critical learning from the dramatic story of Lombardy and many other countries about this first Covid-19 wave, is simply that the lockdown was decreed too late to prevent the overwhelming of health resources. Notably, this occurred despite the evidence from China, despite timely alerts from epidemiologists and mathematical modellers, despite the existence of detailed pandemic plans and instructions developed since more than a decade ago in most western countries but seemingly forgotten (ISS 2007) and, especially, despite the tragic story of Lombardy itself was below every ones’ eyes acting as a formidable advise and effectively communicated (Remuzzi and Remuzzi 2020).

A question is therefore why, in many situations, the lockdown declaration arrived so lately.

In relation to this, a point is whether a delaying role might be played by strategic interactions at the highest political level, besides other factors such as: i) the lack of awareness and understanding among politicians, ii) the widespread circulation of incorrect information and fake news about Covid-19 seriousness. Indeed, there is strong evidence that wild political discussions on the opportunity to declare the lockdown, mainly related to its potential economic damages, continued to occur for long time despite full evidence of sustained transmission. Instances are, for Italy, the incomprehensible choice by the Lombardy local government not to declare hotspots in the Bergamo province, that eventually resulted the most devastated Italian area, despite evidence of a much more serious ongoing epidemic compared to already declared hotspots in Lombardy itself. Other two remarkable examples are the following: the initial decision by the UK prime minister to choose the so called “do-nothing-for-herd-immunity” solution (Stewart et al,, 2020), and the long waiting and multiple hesitations as well as contradictions by French president Macron and his government (Margul 2020; Lexpress and AFP, 2020; Tendance Ouest, 2020; LFI, 2020). All these situations rapidly ended when the growth of serious Covid19 cases proved so large that the lockdown declaration was unavoidable and, seemingly, approved by most stakeholders, though at the price of deteriorated control conditions. Contradictions of all types also characterised the statements by opposition parties in Italy (Dongo, 2020; Mello, 2020) and elsewhere

Here, we report a model-based analysis of the role played by political discussions in delaying the lockdown declaration and consequently in amplifying the epidemic outcome. This is done by a simple game-theoretic framework with three players, the Government, the opposition parties, and the economic lobbies, to represent the context of the political debate on Covid19 in several European countries during the period between the first evidence of pandemic risk and the lockdown declaration. This period is divided into two sub-periods: a pre-epidemic phase, where transmission is not sustained yet and payoffs are fully exogenous, and an epidemic phase, spanning between the onset of sustained transmission and the lockdown declaration, where epidemic costs enter the agents’ payoffs. In particular, we consider a number of situations, depending on the degree of political “responsibility” adopted by the government and the opposition towards their citizens in regard of the epidemic, as reflected in their payoffs.

The paper is organised as follows. In section 2, we present the modelling framework. Section three, presents the key models and results. Concluding remarks follow in the Discussion.

## 2. Model: general ideas

We consider the strategic interaction between three players, namely the Government, its political opposition, and the economic lobbies, during the period prior to the lockdown declaration. The game is as a perfect and complete information one i.e., each player knows both the strategies and the corresponding payoffs of the whole game, though we will consider also the effects of partial information. The game consists in a sequence of one-shot repetitions. Citizens/voters are not explicitly included in view of their limited power in emergency circumstances. However, their preferences are implicitly kept into account by all players, which fear consensus losses. The pre-lockdown period is subdivided into two phases. The first one is the “pre-epidemic” phase, dealing with the waiting-time period between the raising pandemic alert (following, say, the news from the epidemic in China) and the first evidence of sustained indigenous transmission, which we identify as time t=0. The second one, which we will refer to as the true “epidemic” phase, spans between the first evidence of local sustained transmission and the lockdown declaration. In the case of Italy, the pre-epidemic phase ended at 20 february 2020, while the lockdown was gradually implemented between March 5^th^ and March 22nd.

Each player has two possible strategies. The government can decide between implementing either a “timely and harsh lockdown” (strategy “H”) or a milder policy (M). Policy H aims at bringing R_0_ below one and driving the epidemic towards suppression (Ferguson et al 2020, Flaxman et al 2020), by communicating to the population the benefit of accepting such a choice with the resulting economic and social costs. The milder policy (M) which is able to mitigate the epidemic impact while implying less severe economic restrictions, and as such is appealing to economic lobbies. Instances of such a “milder” policy can be considered also the “waiting for events” policy that, arguably, has been adopted in several countries before proceeding with late lockdown.

As for the opposition parties, they can either decide to cooperate with the government by encouraging it (E), or not to cooperate for reasons purely of political consensus, by criticizing (N) the government’s actions towards the emergence. As for the lobby, it can decide whether to encourage the government policy (E) or to criticize it (N).

As a next step, we define three levels of “political responsibility” of political actors towards citizenship. A government is said to be “fully” responsible if it decides to implement the best policy for the society, which is postulated to be H (Ferguson et al. 2020), without caring about consensus. In other words, a government is responsible if playing H is his dominant strategy, i.e. if H is his best reaction regardless of the strategies played by other players. A government is “quite” responsible if neither H nor M are dominant strategies, i.e. if its best reaction is H when the opposition plays E while it is M if the oppositions play N. Finally, a government is “irresponsible” if playing M is its dominant strategy. On the other hand, the opposition can be either (i) irresponsible, if playing N is its dominant strategy, (ii) quite responsible, if its best reaction to H is E and its best reaction to M is N, (iii) (fully) responsible if it is quite responsible and (H,E) makes her equal or better off with respect to (M,N).

As the lobby does not have political responsibility, we do not require for the lobby to be as “responsible” as the other two players. This is reflected in the fact that at epidemic onset, playing E is their best reaction to policy M and playing N is their best reaction to policy H. However, we consider also the case of a quite responsible lobby, i.e. one strictly preferring to encourage policy H rather than M. Instead, we do not consider the case of a fully responsible lobby, since the best reaction to the harsh policy at any individual shot of the game is always to play N.^1^

The analysis goes as follows. First, we consider the strategic interactions between the three actors depending on the mutual degree of responsibility of the government and of the political opposition in an “abstract” setting where payoffs are fully exogenous (i.e., independent of the epidemic incidence). This yields to nine different subcases that are of interest because each one might represent a “real” political context of the pre-epidemic phase, where the exogeneity of payoffs essentially reflects agents’ priori opinions about epidemic risks combined with general objectives belonging to the political debate during ordinary periods as e.g., maintaining political consensus high. In this respect, the analysis of the pre-epidemic phase is critical to identify the role that the degree of political responsibility of the government and the opposition plays on the possibility that the system is prepared to implementing - or not - the lockdown (policy H) as soon as evidence of sustained transmission becomes available.

Subsequently, we move to consider the strategic interaction during the epidemic phase (time t>0), starting when evidence of local sustained transmission is established. During the epidemic phase, all players will necessarily include the perceived (direct and indirect) costs of the epidemic into their payoffs, as explained below.

## 3. Modelling key subcases and results

We discuss more in depth a few main subcases, by distinguishing between the pre-epidemic phase and the true epidemic phase.

### 3.1 The pre-epidemic phase

Our main result (*Table 1*) describes the outcome of the nine games arising from the possible mutual “degrees of responsibility” of the government and the opposition, in terms of the Government’s propensity to declare immediate lockdown (“Yes”), or not (“No”) in the event of onset of sustained transmission. Immediate lockdown always occurs if the government is fully responsible (regardless of the attitude of the opposition) and never occurs for an irresponsible government or for an irresponsible opposition (even when the Government is quite responsible). Moreover, there are two situations where the outcome is ambiguous, both occurring when the Government is only quite responsible.

**Table 1.** The pre-epidemic phase. Outcome of the strategic interaction in terms of propensity to immediate lockdown in the event of onset of Covid-19 sustained transmission. The outcome is given for each of the nine combinations of the possible degrees of responsibility of the two political players (Government vs opposition).

| *Oppositions* | *Fully* | *Quite* | *Low* |
| --- | --- | --- | --- |
| *Government* |  |  |  |
| *Fully* | Yes | Yes | Yes |
| *Quite* | Yes/No | Yes/No | No |
| *Low* | No | No | No |

#### 3.1.1 A “responsible” government against an “irresponsible” opposition

The first best (A) for the government is implementing the policy H without critiques from the opposition, because this leads to the optimal health outcome (epidemic suppression) without credibility losses. The second best (B) is when it implements the mild policy (M) without critiques (especially from the opposition), because it anyhow reaches some results on epidemic mitigation credibility losses. The third best (C) is when it implements policy H facing the attack of, at least, the opposition. Of course, the worst outcome (D) occurs when it adopts the mild policy, and is publicly blamed for it. Clearly, A *>* B *>* C *>* D. The first best for the oppositions is when the government chooses the mild policy and it is publicly blamed, because in this way they feel to sharply gain consensus. The second best is when the epidemic is suppressed and they cooperated with the government, achieving the optimal health result without credibility losses. The third best is when the government implements policy H and the oppositions criticise it (this can become the second best if also the lobby attacks the government because the epidemic is suppressed and at the same time the lobby starts supporting the oppositions). The worst case occurs when the government chooses mitigation, with a poorer health outcome, and the opposition encouraged it, therefore missing the possibility to gain a consensus advantage with respect to the government. The third and fourth best switches if the lobby encourages the harsh policy of the government, because in this way the loss of credibility is more likely to occur for the opposition refusing to encourage epidemic suppression. The lobby’s first best is when the government implements M and the lobby encourages it, because we assume that the lobby’s credibility is irrelevant outside its own members. The second best is when the government implements policy M and the lobby criticises it. The third best is when policy H is implemented and the lobby criticises it, because at least the lobby is coherent with respect to its members interests. The worst case is when H is implemented and the lobby encouraged it, because the lobby will suffer both economic and credibility losses.

To find the Nash equilibrium of this game, we build the reaction of the opposition and the lobby with respect to the possible strategies (H,M) of the government (*Table* 2). The resulting payoffs are reported in each cell according to the following order: government, opposition, lobby.

**Table 2.**
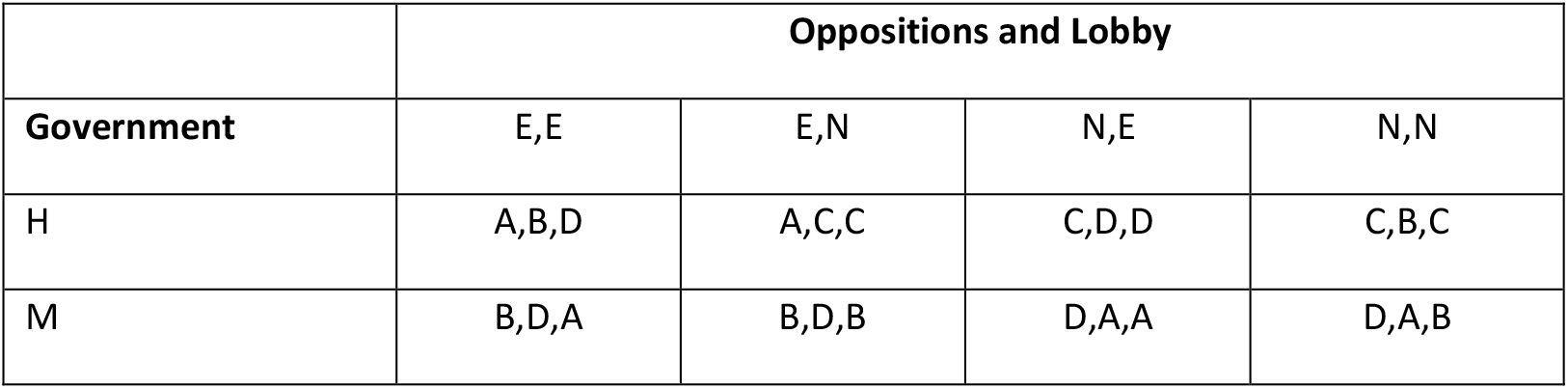
The case of a “responsible” government against an “irresponsible” opposition: strategic interaction between lobby and opposition vs the government.

By applying iteratively the strict dominance criterion, starting from whichever player, the best reaction to policy H is then to discredit it. Pairwise, in case the government implements policy M, the best reaction is when oppositions criticize it, while the lobby encourages it.

From now on, we will neglect the lobby, since we know they will criticize a priori policy H and instead will promote policy M.

Since the game is one with complete information, the government is simply left to choose between implementing H and getting blamed for it (obtaining a payoff of C), or implementing M and getting blamed by the opposition for it (obtaining a payoff of D). The strict dominance criterion ensures that the government will eventually implement H, though facing blaming from the other players.

Notably, the Nash equilibrium is the third best for both the lobby and the government, and the second best for the opposition, which is the player which is better off at the end of the game. Also, as epidemic suppression is the first best from the societal standpoint, it happens that citizens are the true winner of the game despite they do not play it, while the government and the lobby, that should be the strongest players, end up worse off than the others.

To sum up, this model relies on a narrow-minded opposition, only interested in criticizing a priori the government, and therefore preferring the government be mistaken, thereby increasing the epidemic burden, just to increase its consensus, rather than facing a government suppressing the epidemic, despite they will also benefit from the suppression.

As a final remark, note that all the configurations are Pareto optimal, though implementing policy H implies intuitively a better overall wellness if we would include the whole society. Note finally that a Stackelberg game where the government moved first would lead to the same results.

#### 3.1.2 A quite responsible government vs a fully responsible opposition

Here we consider the case where the government is only “quite” responsible i.e., it mostly fears to be blamed and therefore has the objective to get at least some support from other players. Thus, even if it would prefer to implement the H being supported by the opposition, it is ready to switch to M and being supported by the lobby if the opposition decides to criticize its actions. On the other hand, we consider a responsible opposition which would like to support policy H or at least to criticize the mild one (though the ordering of these two alternatives can be switched without altering the main content of the game).

This results in a coordination (“stag hunt”) game (Table 2) where the two pure Nash equilibria are (H,E) and (M,N), and the mixed strategy equilibrium is characterized by p=(C-D)/(A-B+C-D) and q=(B-D)/(A+B-C-D), where p is the probability that the oppositions play E and q is the probability that the government plays H.

**Table 2.** *The case of a “quite” responsible government against a “responsible” opposition: strategic interaction between lobby and opposition vs the government*.

|  | Oppositions and lobby |  |
| --- | --- | --- |
| Government | E | N |
| H | A,A,C | D,C,C |
| M | B,D,A | C,B,A |

First of all, note that the equilibrium (H,E) Pareto dominates all the others (disregarding the lobby). Nevertheless, the outcome might also be (M,N), resulting in the first best for the lobby but in a bad outcome for the rest of the society, since the lockdown would be delayed and the other players would be better off by cooperating. This may happen despite the opposition is more responsible, the government is still quite responsible, and the lobby seems to have no strategic relevance. The reason is that the lobby is pivotal in the consensus process. This can cause a lack of trust between the other two players, inasmuch as they are not sure whether the opponent will trust them, and cooperating while the other defects will lead the co-operator to get the worst payoff D. In other words, if the government implements policy H and the opposition criticise it, the government will fear to lose consensus and also the lobby’s support. On the other hand, if the oppositions publicly support the government but the government decides to implement the mild policy, then they will both loose consensus and the lobby’s support. Actually the result of the game depends upon the subjective probability that each players assign to the other choosing to cooperate.

Technically speaking, the two pure strategy Nash equilibria are stable, while the mixed strategy equilibrium is unstable: it is sufficient that the players think the opponent will cooperate with a probability slightly higher(lower) than the mixed strategy equilibrium’s one to converge immediately towards full cooperation(defection).

Overall, the final outcome will depend on the relative difference between the values *A, B, C, D*, which are a measure of the risk arising from trusting the opponent. In a more realistic version of the game, players’ believes on the opponent’s action may depend on the past history, i.e. how many times the government and the oppositions have been able to cooperate.

#### 3.1.3. A quite responsible government vs a quite responsible opposition

This case is found by simply switching A and B for the oppositions in Table 2. The result is again ambiguous: (H,E) and (M,N) are pure strategy Nash equilibria, and there exists also a mixed strategy equilibrium (*q′, p′*) with *q′ > p′*. Therefore, an even higher subjective probability (compared to the case of sub-section 3.1.2), is required to lead the oppositions to cooperate with the government.

### 3.2 The epidemic phase

Decisions and payoffs from the pre-epidemic phase (section 3.1), are inherited by players at onset of the epidemic phase, which starts once evidence of sustained transmission becomes available. After this moment, which we identify by time t=0, epidemic data enter the agents’ payoffs, which are now taken as the sum of a fixed component, given by the payoffs from the pre-epidemic phase, plus a component having negative sign, reflecting the agents’ perceived costs of the epidemic. The latter costs are defined as the sum of the direct health cost of the epidemic, and of its indirect, mostly economic, cost related to the effects of the deployed interventions on economic activity. Direct costs are assumed to be incidence-dependent that is, to depend on some appropriate index *I_t_* of the epidemic incidence, for example an average of the observed incidences of Covid-19 confirmed cases and Covid-19 deaths. Indirect costs are assumed to depend linearly on the duration of the lockdown through a constant coefficient reflecting the GDP loss per day. We assume that in the epidemic phase index *I_t_* obeys exponential growth according to a Covid-19 transmission model parametrized by data from the epidemic early phase in Europe (appendix). Direct and indirect costs, are weighted differently by players according to each specific player’s features. Let functions *F*(*I_t_*) (*f*(*I*_t_)) and *G*(*D_t_*) (*g*(*D*_t_)) denote the epidemic time-dependent direct cost resulting from policy M (H) when the incidence is *I_t_*, and the lockdown duration is *D_t_*. As stated above we set: *G*(*D_t_*) *= G*_0_ *· D_t_* and *g*(*D_t_*) = *g*_0_ *· D_t_*. Instead, functions *F*(*I_t_*) (*f* (*I_t_*) are nonlinear to reflect the dramatically different impact of the two possible policy actions H,M in terms of epidemic control, as underlined first in Ferguson et al (2020). Just for sake of illustrations we take *F* increasing and convex in *I_t_*,and *f* increasing and concave (*F′ >* 0, *F″ >* 0*, f′ >* 0 *, f″ <* 0 where derivatives are taken with respect to *I_t_*.The implementation of policy H at a certain time point *T* abruptly reduces the values of *F_t_* to *f_t_* but expands the value of *g_t_* to *G_t_*. The three players weight differently the two cost items, with *α, γ,β*, representing the weights of direct costs for the government, the oppositionand the lobby, respectively. We assume *α > β,γ > β*. The general structure of payoffs during the epidemic phase is described in Table 3.

**Table 3.** The general structure of agents’ payoffs during the epidemic phase. Payoffs in each cell are ordered as follows: Government (first row), opposition (second row), lobby (third row).

|  | Oppositions |  |
| --- | --- | --- |
| Government | E | N |
| H | $A - \alpha f(I_t) - (1 - \alpha)G(D_t),$<br>$B - \gamma f(I_t) - (1 - \gamma)G(D_t),$<br>$C - \beta f(I_t) - (1 - \beta)G(D_t)$ | $D - \alpha f(I_t) - (1 - \alpha)G(D_t),$<br>$C - \gamma f(I_t) - (1 - \gamma)G(D_t),$<br>$C - \beta f(I_t) - (1 - \beta)G(D_t)$ |
| M | $B - \alpha F(I_t) - (1 - \alpha)g(D_t),$<br>$D - \gamma F(I_t) - (1 - \gamma)g(D_t),$<br>$A - \beta F(I_t) - (1 - \beta)g(D_t)$ | $C - \alpha F(I_t) - (1 - \alpha)g(D_t),$<br>$A - \gamma F(I_t) - (1 - \gamma)g(D_t),$<br>$A - \beta F(I_t) - (1 - \beta)g(D_t)$ |

The analysis of the game during the epidemic phase focuses on the role played by the degrees of responsibility of the government and the opposition, as represented by the payoffs inherited from the first phase, and by the relative weights *α, γ, β* agents pay to direct vs indirect epidemic costs. Instead, cost functions *F_t_, f_t_, G_t_, g_t_*, are not agents-specific. Effects of agents’ myopia, as represented by differential delays with which costs are perceived by the different agents, are discussed in the appendix.

#### 3.2.1 Delayed lockdown declaration: mutual role of the government and the opposition

We assume that when the lockdown implementation occurs instantaneously after declaration, by abruptly halting non-critical activities, and that no indirect costs arise from the pre-lockdown phase (*G = g*=0). Note preliminary that if *α =* 0, i.e., the case of a purely “mean” government not including direct epidemic cost in its payoffs, the lockdown will never be declared and even the most responsible opposition would be worthless. Instead, if *α >* 0, H policy will always be implemented sooner or later, regardless of the initial position, due to the disproportionate growth of direct costs. Obviously, for situations in Table 1 showing propensity to lockdown already from the pre-epidemic phase, the lockdown declaration will occur without delay after onset of sustained transmission. For cases leading to a positive lockdown delay, it is sufficient to consider only one case from Table 1, as all remaining ones will straightforwardly follow.

Based on previous remarks, we focus on the more interesting case where at onset of sustained transmission both players, though quite responsible, were in the Nash (stable) equilibrium (M,N) (discussed in section 3.1.3). In that equilibrium, the government has not imposed the lockdown yet, thereby being supported by the lobby, while the oppositions is criticizing by calling for immediate lockdown.

The subsequent events depend on the payoff switch of the government and the opposition, which will occur for:

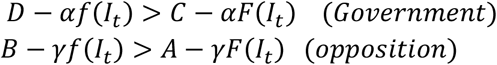

that is, for

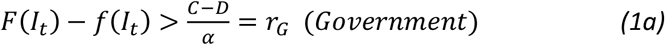

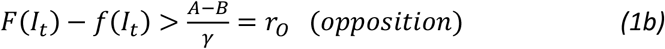

where the difference *F* (*I_t_*) *− f* (*I_t_*), represents the net additional (direct) cost of policy M with respect to policy H. Therefore, if the government is, overall, more responsible than the opposition (*r_G_* < *r*_0_), the first agent leaving the (M,N) configuration (which will not be a Nash equilibrium anymore) will be the government. More precisely, there exists a time t=T at which the government will adopt strategy H, by consequently declaring the lockdown. On the other hand, the opposition will suddenly react by playing E, even if it will remain in a suboptimal configuration up to the shots in which (1*b*) will be fulfilled. Pairwise, the lobby will switch when *F*(*I_t_*) *− f* (*I_t_*) *>* (*A − C*)*β*. That means that there are times T_O_, T_L_ (T_L_ > T_O_ > T), where both the opposition and the lobby will adjust to their optimal configurations, though this has no direct impact on the actions they undertake. Obviously, other things being equal, the lower *r_G_* that is, the lower the degree of responsibility of the government, i.e., the later the lockdown declaration will occur (and vice-versa).

This is illustrated in Figure 1, showing an initial phase where the fixed components of payoffs inherited from the pre-epidemic phase dominate the incidence-dependent components. However, as the epidemic grows, function *F* overwhelms function *f* by more than the difference between the fixed components, so that a switch in the ranking of payoff occurs for all players. Given the assumed ranking in terms of degrees of responsibility, the Government switches first, declaring the lockdown with a delay of about seven days after evidence of sustained transmission.

**Figure 1.**
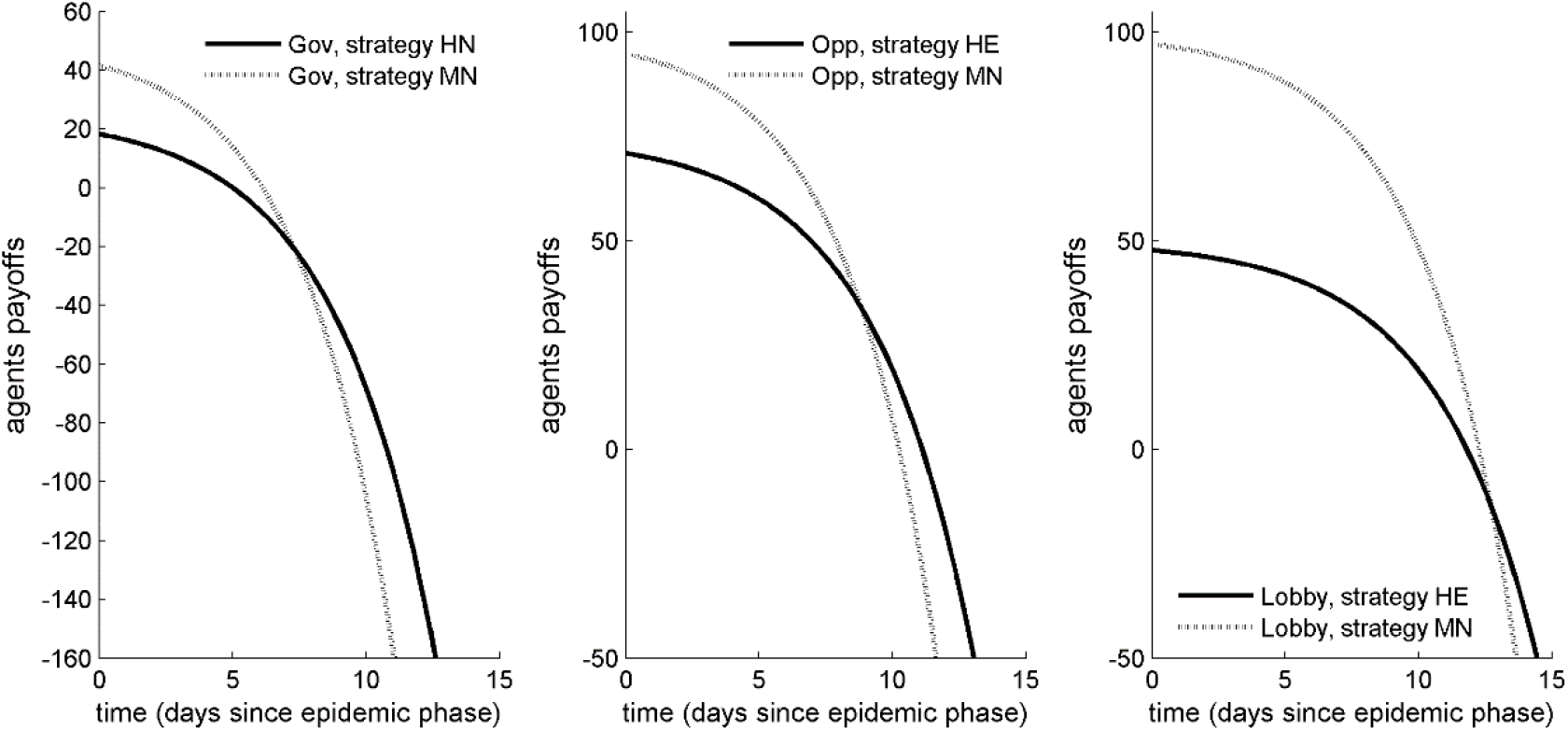
The epidemic phase: trends in the payoff of the three players. Left panel: Government. Central panel: opposition. Right panel: lobby. Fixed payoffs from the pre-epidemic phase: A=100, B=75, C=50, D=25; weights of direct epidemic costs: *α =* 0.75 *,γ =* 0.45*,β =* 0.25, implying *r_G_ =* 33.3*,r_O_ =* 55.6. The lockdown declaration by the Government at time T (around day 7), occurs at the time when, during the phase of epidemic exponential growth, the payoff of strategy HN overtakes the corresponding payoff of strategy MN.

#### 3.2.2 Anticipated lockdown resulting from a fully responsible behaviour of the opposition

The fact, shown in the previous section, that the government decides at time shot *T* to implement the lockdown without caring about cooperating with the opposition may imply that, if coordination were possible, the whole society could benefit from an earlier lockdown. Indeed, an opposition overall “more responsible” than the government (*r*_0_ < *r_G_*) will experience the payoff switch at time shot T_O_ < T. At this stage, the opposition will prefer to cooperate with the government rather than to stick in the non-cooperative Nash equilibrium. In the illustration of Figure 2, this coordination will allow the lockdown to be anticipated at time T_L_=2.5 days, thereby “saving” almost two further doubling times of the epidemic. Other things being equal, this will occur if the opposition is responsible enough, that is if in the pre-epidemic phase the opposition cares epidemic containment almost like preserving consensus. Nonetheless, enacting cooperation will require to solve the resulting coordination issues.

**Figure 2.**
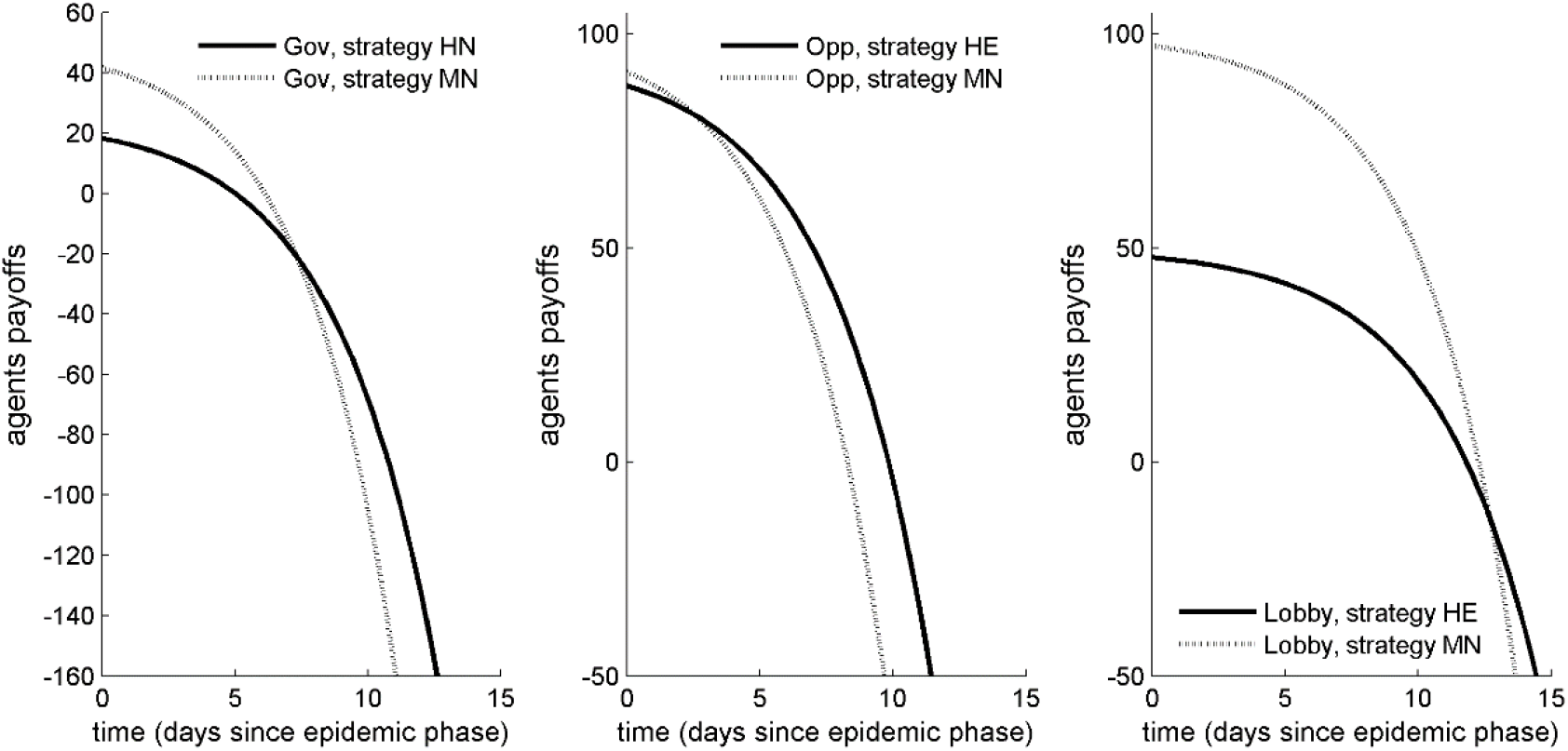
The epidemic phase: trends in the payoff of the three players. Left panel: Government. Central panel: opposition. Right panel: lobby. Fixed payoffs from the pre-epidemic phase: A=100, B=90, C=50, D=25. *α =* 0.75,*γ =* 0.80*, β =* 0.25, *r_G_ =* 33.3, *r*_0_ *=* 6.25. The presence of a “responsible” opposition (the payoff of strategy HE overtakes the corresponding payoff of strategy MN at day 2.5), allows the possibility to anticipate the lockdown compared to the moment (day 7), when the Government would proceed with the lockdown declaration.

#### 3.2.3 Conditions for cooperation between government and opposition allowing un-delayed lockdown

Note that in the extreme case of a fully responsible opposition (*B* ≥ *A*), i.e., caring societal health equal or more than political consensus, the lockdown can in principle occur without delay i.e., for *T = 0*. This would be the first best for the society as a whole and for all players but the lobby (which however should be in its first best too, given that we are disregarding the fact that the earlier the lockdown the shorter its duration). In this way, at time *τ* the ranking of cells is the same as depicted in Table 2, and the outcome will be (H,E) provided that both the government and the oppositions assign each other a subjective probability of cooperating at least equal to the probabilities computable resorting to the mixed strategy.

We note that this cooperation outcome, though in principle critical, is not as straightforward as it might seem, because it requires to solve the communication problem underlying coordination. Two problems arise.

First of all, if agents are myopic (see the specific section in the appendix) in the sense that upgraded information on the state of the epidemic appears with a lag s (adding to the objective delays intrinsic to Covid-19 dynamics), and *s > T − T_0_*, then earlier lockdown becomes impossible, though it would have been optimal for everyone.

Second, it can happen that the subjective probability that each player assigns to the possibility that the other player will cooperate is low, e.g. because *s* is large.

There are two possibilities, both implying sending a message to the other player. Let focus on shot *T_0_* = *τ* and denote by *δ_G_* and *δ_0_* the discount rates of the government and the opposition, respectively. Since the coordinative game is repeated *T* − *τ* times (up to the point in which playing H becomes the dominant strategy for the government), one or both player could cooperate in the first shot, letting the other understand that he will cooperate. Let us introduce the abridged notation *a_t_ = A − αf* (*I_t_*), *d_t_ = D − α f* (*I_t_*) and so on for the government and 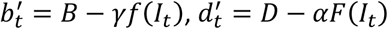 and so on for the opposition. The payoffs emerging from sending this kind of message is:

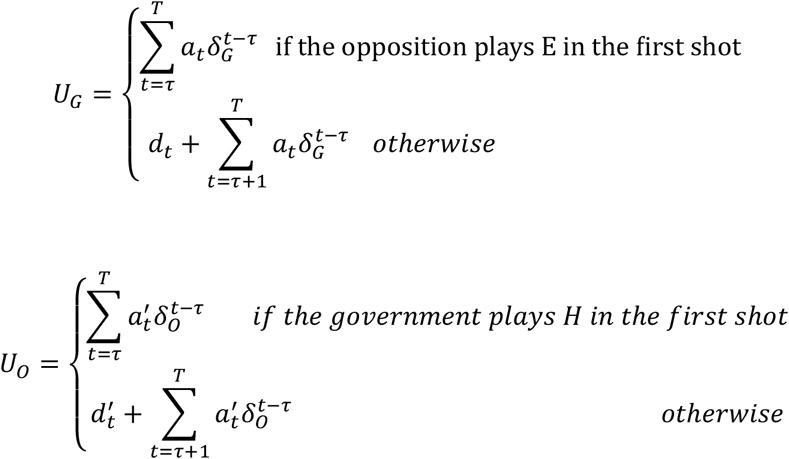

On the other hand, waiting up to shot *T* before cooperating leads to:

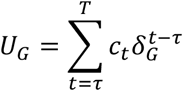

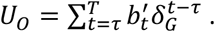

Therefore, the government will play strategy H already from the first shot if:

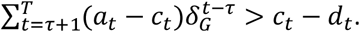

(Actually the threshold is even lower since we consider only the worst case, i.e. the other player does not send any message).

On the other hand, the opposition will play E from the first shot if:

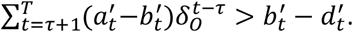

If the first condition holds, the lockdown will start at shot *τ*, if only the second holds it will start at shot *τ+*1.

Another kind of communication could be achieved if the government had the possibility to publicly speak about the opposition as well. Consider a repeated two stage game where the government decides whether to say that the opposition is very responsible or to say that it is irresponsible before he implements the policy of time *t*. In the second stage, players simultaneously play the original one shot game. If the government communicates that the opposition is doing well and then they do not coordinate, the government will get *c_t_ − r*, where *r* is a reputational cost, while if instead they coordinate the government(oppositions) will get 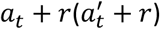. Simple analysis leads us to conclude that if the government says the opposition is doing well, by forward induction the opposition will play E, and the government will play H. This might be an effective solution if the conditions of the repeated game above are not satisfied for the government.

### 3.3 Epidemiological implications

#### 3.4.1 Delayed lockdown

The effects of lockdown timing against Covid-19 have been discussed in a number of papers (e.g., Walker et al. 2020). We therefore only report results of interest for the present work. We use a simple homogeneous mixing epidemiological model (appendix) parametrised with a range of data from the Italian Covid-19 experience (Cereda et al 2020, Guzzetta et al 2020, Riccardo et al 2020). The model population is chosen to mirror the population above 60 years (about 20 million) resident in Italy at January 1st, 2020 (UN 2020), where most Covid serious morbidity and mortality occurs. The basic reproduction number is set to *R*_0_ = 3.25 to produce a doubling time of about 2.75 days. We consider first the case of a harsh and sudden lockdown abruptly bringing *R*_0_ to the subcritical level *R_t_* = 0.8, and keeping it constant thereafter (essentially until epidemic suppression i.e., disregarding non-health costs). Figure 3 compares the effects of two different scenarios i.e., i) an *early lockdown* (central panel), declared without delay by a fully responsible government (in the sense of Table 1) as soon as the technical committee indicate the urgence to do so, and (ii) a *late lockdown* declared with one week delay, according to the right panel of Figure 1. The former is declared and implemented when the number of hospitalised cases has reached the threshold level of 500 (note, just for comparison, that in Italy the national lockdown was declared on March 11^th^ when the number of Covid-19 confirmed hospitalised cases was in the range of 6000), while the latter is implemented with a seven days delay, when all epidemic markers have grown, given the exponential increase of the epidemic curve, by a factor *exp*(*r ·* 7) *=* 5.8, where r is the real time growth rate of the initial exponential phase (for reference, the left panel reports also the case of a free, uncontrolled epidemic). Figure 3 also reports a “100-hospitalizations line”, arbitrarily chosen as a marker of lockdown success after which unlocking can be considered.

**Figure 3.**
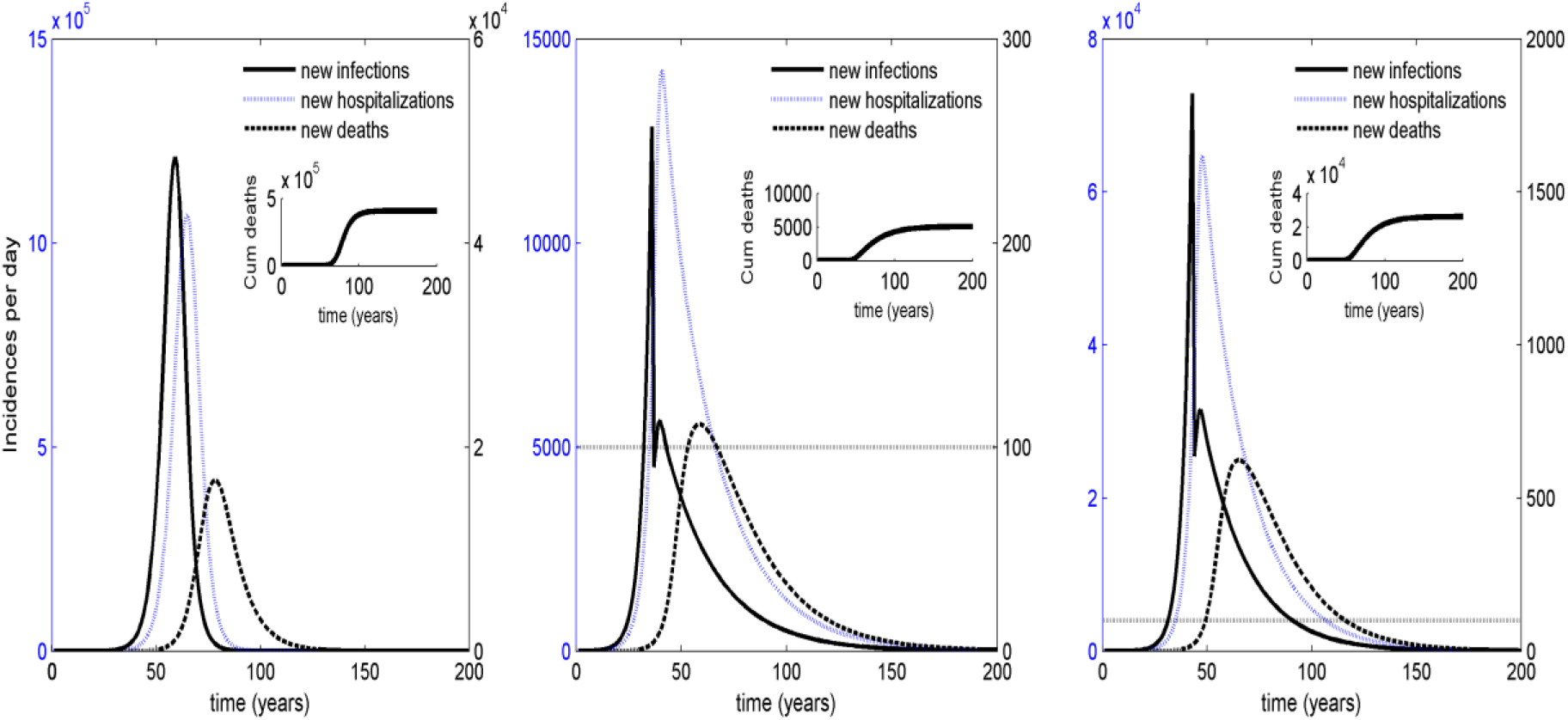
Effects of delayed lockdown. Left panel: the case of a free epidemic, reported for reference. Central panel: the case of an abrupt *early lockdown* initiated when the threshold of 500 hospitalised cases has been reached. Right panel: the case of an abrupt *late lockdown* initiated – due to strategic interactions – with a seven days delay. Left axis: new infections per day. Right axis: new hospitalizations and deaths per day. The graph also reports the 100-hospitalization line.

The two most straightforward implications of the delayed lockdown are the dramatic expansion of the cumulative mortality burden, which increases by more than five times, and the dramatic extension of the duration of the lockdown phase necessary to achieve the desired “100-hospitalised” control target. The latter increases from slightly more than one month up to about three months, making the duration of the lockdown essentially unsustainable for whatever economic system.

Notably, the results presented here are robust to a number of sensitivity factors such as e.g., the lockdown marker (say, using ICU occupied beds, or deaths, instead of hospitalizations) or the lockdown threshold. Moreover, they under-estimate the true outcome as they do not include the possibility that the delayed lockdown brings to saturation the hospital system (which we rule out for early lockdown), thereby further amplifying mortality.

#### 3.4.2. Post-emergency re-opening at non-negligible epidemic levels

As remarked above, given specific control targets, a late lockdown will have to last much longer compared to an early one, becoming potentially unsustainable for the economic system. On the other hand, the success in epidemic control allowed by the lockdown will eventually cause the epidemic curve(s) to downturn, thereby down-turning direct costs, while indirect costs continue to increase. This will, sooner or later, lead to a payoff re-switch. The payoff re-switch will force the Government switching back to strategy M, possibly declaring the end of the lockdown. This is illustrated in Figure 4, under the epidemic conditions of the late lockdown of Figure 3. Figure 4 considers two different scenarios in terms of different evaluations of the indirect cost of the epidemic, both yielding to the payoff re-switch long before the established control targets have been achieved, potentially hindering the benefit of the lockdown.

**Figure 4.**
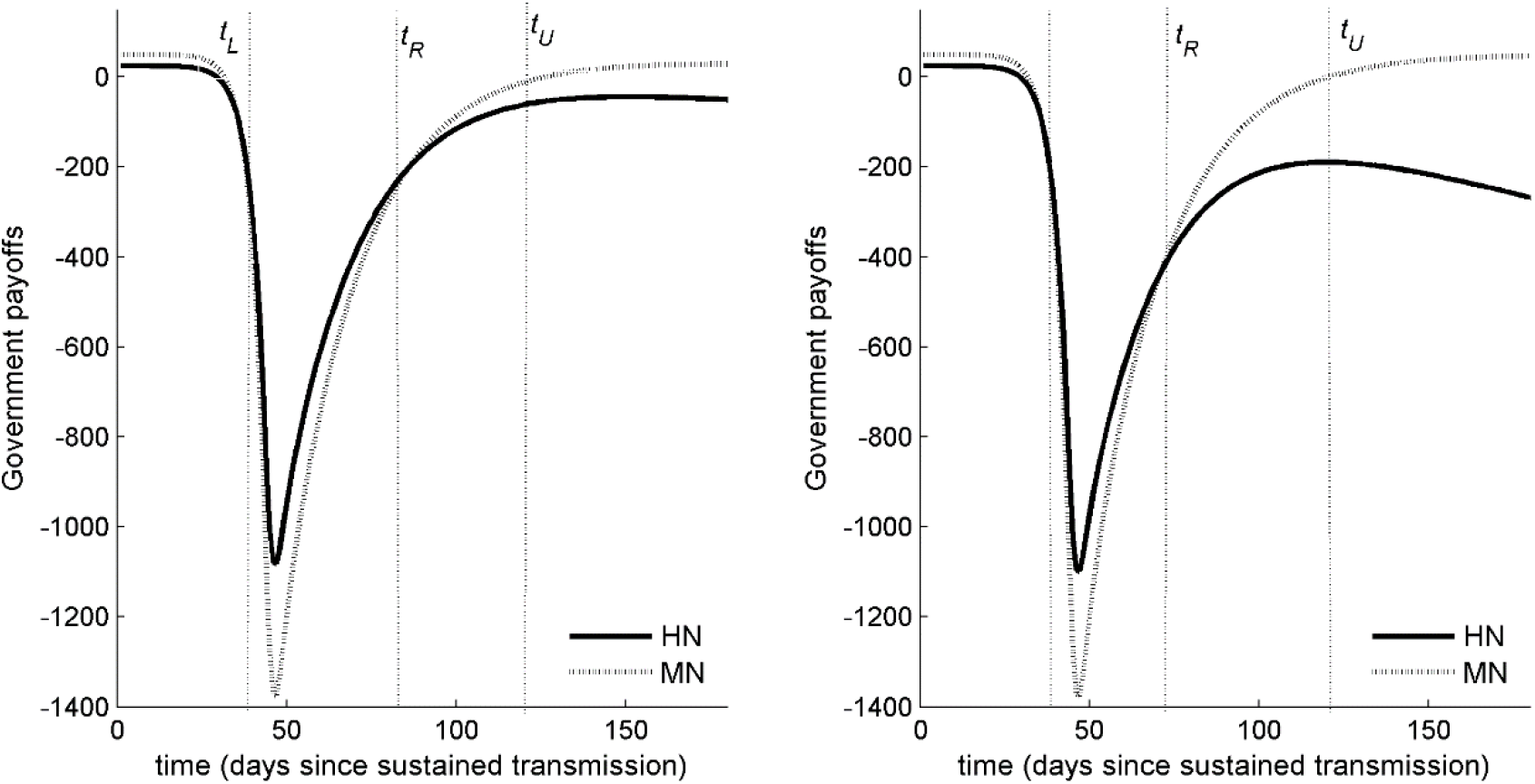
Lockdown declaration at time *t_L_ = T* resulting from the government first payoff switch and subsequent unlocking due to the government payoff re-switch at time *t_R_*, under the epidemic conditions of the right panel in Figure 3. Left panel: low indirect cost of the epidemic (*G*_0_ “low”), Left panel: high indirect cost of the epidemic (*G*_0_ *“*high”). Notably, *T_R_* occurs long before the appropriate control targets for unlocking (i.e., the achievement of the 100 hospitalizations line in the right panel of Figure 3) have been reached at time *t_U_* (*T_U_* about 120 in Figure 3).

This suggests a vicious circle where in a first phase of the epidemic mild degrees of responsibility of the government and the opposition (and lobby pressures) forced the lockdown declaration to be delayed, eventually needing a longer lockdown (due to the much worse epidemic outcome, despite lockdown). But the longer lockdown will unavoidably increase its economic impact, thereby motivating lobbies and low-responsible political parties to. Earlier shout for the need to unlock unlocking i.e., in epidemiological terms, faster re-bringing *R_t_* above unit long before the epidemic has reached adequate levels of suppression, will speed up the epidemic restart, thereby accelerating the need for further lockdown.

Such a vicious circle seems to have occurred in many places during the first Covid-19 wave e.g., at least in Lombardy, Spain the UK and the US.

## 4. Discussion and concluding remarks: the earlier-the harsh, the better (for all)

The short story of Covid-19 has shown a range of fundamental issues in relation to the available options of epidemic control. Ruling out – unless alternative choices are unavailable - the “herd immunity”, or “doing nothing” solution, initially invoked by the UK Prime Minister, the first of these issue is that border surveillance plus containment at onset are difficult due to the presence of a remarkable number of pre-symptomatic, asymptomatic and pauci-symptomatic subjects. If such measures fail, and the epidemic spreads, the only strategy to contain its dramatic burden, protecting fragile people and avoiding hospital collapses, is by resorting to extreme measures, namely early and intensive lockdown. The earlier the lockdown is declared, the lower the negative impact on economic. Indeed, the lockdown duration required for adequate epidemic control, will in this case be minimal.

These facts are undoubtedly clear from the evidence comparing situations of late lockdown e.g., Lombardy Region, against situations, as regions of Central and Southern Italy (Riccardo et al 2020), where the national lockdown declaration came for these regions at a sufficiently early stage of the epidemic.

However, the Lombardy case showed that, if delayed, even a full lockdown cannot prevent a prolonged mortality bulge due to the collapse of the diagnostic chain and hospitals (Remuzzi and Remuzzi, 2020, Ferguson et al 2020). In other words: there is a maximal lockdown date. Notably, the time window for lockdown declaration is short due to the fast timescale of Covid-19 growth characterised by a doubling time of 3 days about (Cereda et al 2020, Guzzetta et al 2020) and to its intrinsic “delays”, namely: i) the presence of a latency period and ii)the time needed for case-confirmation procedures. These delays imply that cases confirmed today result from infections caught even, say, 12 days ago.

In this work, we have used a simple game-theoretic framework to analyse the inertia effects in political decisions that arise due to the strategic interaction between the political actors in such emergency situations, namely the Government, the opposition, and myopic economic lobbies opposing the lockdown due to its presumed economic costs.

Our results are as follows. As a rule, the lockdown will always be declared immediately (i.e., without delay) as sustained transmission arises, only if the government feels to be fully responsible towards all citizens. This situation is unlikely to occur as an outcome of the pre-epidemic phase, due to the fact that the political debate (and payoffs) in this phase will still be biased by evaluations typical of ordinary political phases. Second, unless the government does not attribute any cost to the epidemic, the exponential growth of the epidemic will eventually dominate the agents’ payoffs, so that – sooner or later - the lockdown declaration will always occur. That is, both the government and the opposition will unavoidably be forced by the epidemic to switch towards a higher degree of “responsibility towards citizens” regardless of their initial position. However, this will occur with a delay. There is a further nontrivial situation where the lockdown can be declared with short or no delay, which arises when the political opposition is at least as responsible as the Government since the beginning, or when it becomes such during the epidemic course. This however requires the solution of coordination issue which cannot taken for granted. Finally, the success of the lockdown in containing the epidemic and reducing incidence will unavoidably bring a situation where a payoff re-switch will occur, causing players to return to a situation of lower responsibility. This might cause an untimely unlock of activities, thereby promoting a fast return of R _t_ above threshold. As a consequence, epidemic restart given that the susceptible proportion in the population likely is still very high. This unless the majority of citizens have ’interiorized’ the lockdown lesson and spontaneously maintain for a sufficiently long period a reduction of behaviours at risks of infection. Tis, however, would imply that the behavioural changes of citizens are fully or mildly dependent on the information on the disease prevalence, which is unlikely (Manfredi and d’Onofrio, 2013). Another possible scenario is that the *unlockdown* coincides with external factors possibly reducing the disease transmission, e.g. increase of temperature (Buonomo et al, 2018).

To sum up, our main point here is that the presence of a lower initial degree of players’ political responsibility, jointly with inappropriate evaluation of epidemic cost and pressure enacted by economic lobbies, can be responsible of substantial delays in the timing of the lockdown declaration. But these delays will add to the objective delays that are intrinsic to Covid-19 transmission (latency times and delays related to cases confirmation process), thereby causing a dramatic amplification of epidemic impact, even in the presence of a full lockdown. Just to exemplify in plain language, suppose science suggests that the lockdown should be declare today based on some epidemic marker. This implies that the first evidence of the lockdown success – in terms of a slowing down of the epidemic curve - will be detectable only after say 12 days, during which – given a 3 days doubling time - a 16-fold increase in cases, hospitalizations and deaths is expected to occur. However, if political inertia causes a further 6-9 days delay – that is 2-3 further doubling times - prior to lockdown, the observed growth would bring a 64-128-fold cases increase– a very large epidemic almost surely overwhelming whatever hospital resources might be available.

A further negative impact would be at world-wide scale. Indeed, untimely end of lockdown would be dependent on local dynamics of the disease, thus causing a heterogeneous distribution of the disease dynamics, which potentially could favour the pandemics.

More than this, we would like to pinpoint the pernicious role that mild degrees of responsibility of the government and the opposition, combined with lobby pressures can have (and actually had) on the entire war against Covid.-19. Indeed, these factors can delay the lockdown declaration, eventually implying the need of a longer lockdown (due to the much worse epidemic outcome, despite lockdown). But the longer lockdown will unavoidably increase its overall economic impact. And the economic losses will motivate the lobbies and low-responsible political parties – which caused the lockdown delay - to ask for the need to unlock earlier given the economic damage. Thereby accelerating the need for further lockdown and potentially increasing the economic damage they claim they want to mitigate.

The limitation of our approach, on which we are working, concerns three main areas: i) our model is essentially deterministic, and as such it does not consider sudden stochastic perturbations affecting political decisions (e.g. announcement by EU); ii) our model is at single country scale, which we think it is an accurate modeling for this phase of the Covid-19 pandemics, but for a longer period of time a multi-state version of our models could be important; iii) we did not explore possible interplays between the epidemics time-scale and the disease time-scale

It is worth to note that in the increasingly growing field of behavioural epidemiology of infectious diseases (BEID) (Funk et al 2011, d’Onofrio et al 2012, Manfredi and d’Onofrio 2013, Wang et al 2016) the government actions modulating the citizen behaviour during an epidemics are considered in an elementary way. Indeed, in BEID the emphasis was up to know given to the modeling of the citizens’s behaviour. Here, at the best of our knowledge, we introduce an explicit and *’disease dynamics-dependent’* modelling of the government behaviour via a game-theoretic approach.

Under this light, we may say that our work uses theoretical arguments of BEID to suggest the harmful health impact that has potentially been caused by mean political behaviour during the pre-lockdown phase. This mean behaviour is apparent from the public political debate occurred in many European before epidemic events forced the lockdown. All this, despite the evidence from China and from Lombardy. A full reconstruction of these harmful political behaviour will be important in future research. The role of historians will be fundamental in this scenario where the usually long times-scales of history were hugely accelerated,

The planet will have to coexist with Covid-19 for a long period of time. Countries that chose the “suppression” strategy must be prepared - given the large susceptible proportions remaining – to declare further lockdowns in the future. The present warning about both the timing of locking and unlocking should be made fully clear to policy makers and lobbies, not only to inform their decisions and behaviour, but especially to call them to higher degrees of political responsibility, in order to avoid the tragic mistakes occurred during this first pandemic wave.

## Data Availability

This is a theoretical work. All data used can be found by simply googling them.

## Appendix

### A1. The effects of information asymmetry and myopia

Myopia is a potentially intrinsic characteristic of players’ information set during the “Covid game”. A first component is almost “objective” and related to the natural delays by which cases appearing to the public health system “today” actually represent infections occurred up to 10-15 days before. This component is objective because even if the problem is perfectly known to, first of all, the Government experts, no lockdown is declarable before having observed at least some cases. Further components might be due to the more complete information available to the Government about epidemic trends in view e.g., of the direct support it receives from technical and scientific institutions, namely the National Institute of Health, and the prevailing emergency conditions. The effects of myopia into our simple framework are quite straightforward: the incidence function reveals itself with a time-delay causing further delays in the payoff switch yielding to the lockdown declaration. The worst case is when the myopia also involves the Government decisions because it will severely inflate the lockdown delay. This sounds to have been the case of the UK.

In presence of information asymmetries represented by time delays in the perceptions of the epidemic seriousness, the payoff matrix is represented in the Table below

**Table 4.**
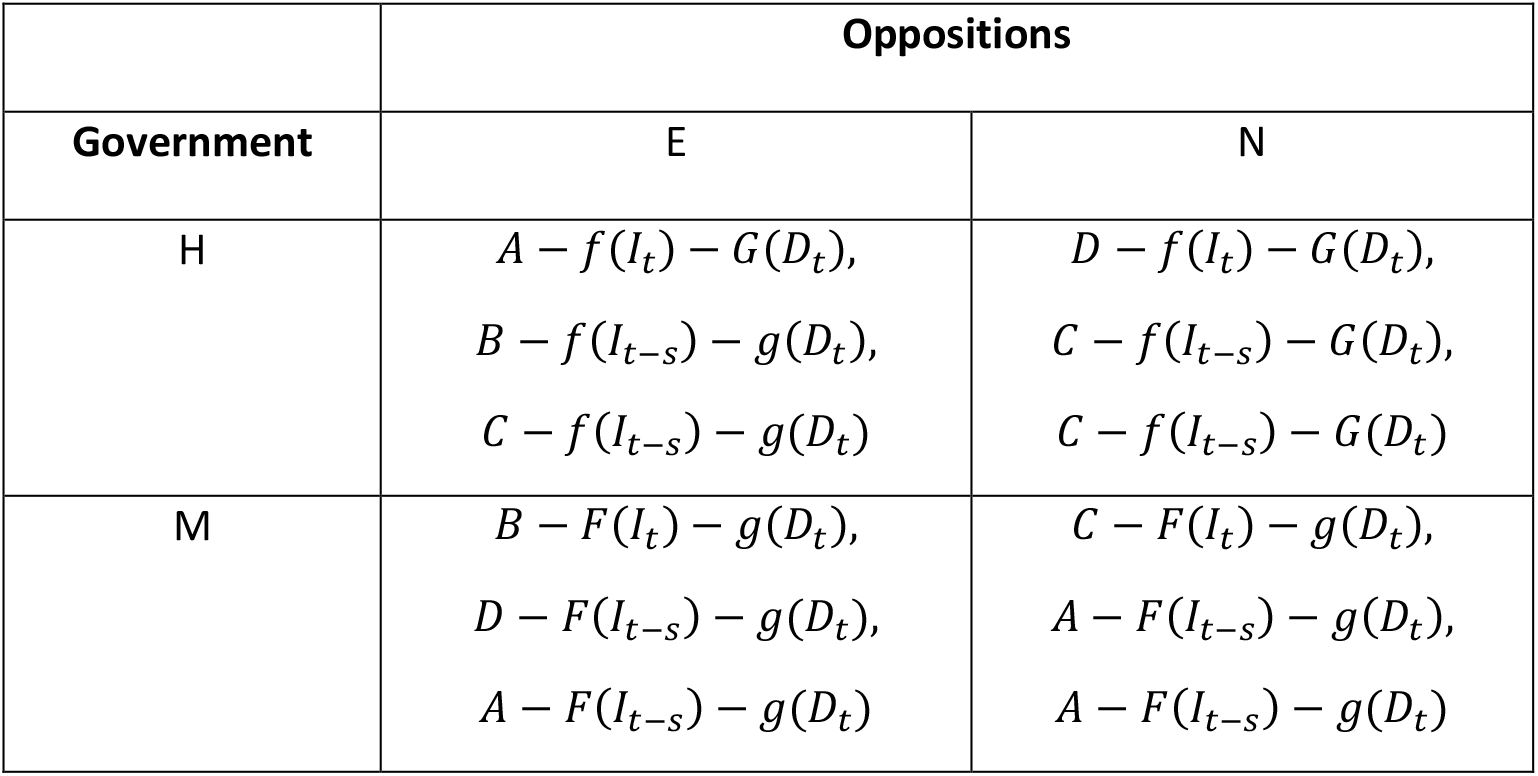
The game with direct inclusion of epidemic dynamics and economic cost.

|  | Oppositions |  |
| --- | --- | --- |
| Government | E | N |
| H | $A - f(I_t) - G(D_t),$<br>$B - f(I_{t-s}) - g(D_t),$<br>$C - f(I_{t-s}) - g(D_t)$ | $D - f(I_t) - G(D_t),$<br>$C - f(I_{t-s}) - G(D_t),$<br>$C - f(I_{t-s}) - G(D_t)$ |
| M | $B - F(I_t) - g(D_t),$<br>$D - F(I_{t-s}) - g(D_t),$<br>$A - F(I_{t-s}) - g(D_t)$ | $C - F(I_t) - g(D_t),$<br>$A - F(I_{t-s}) - g(D_t),$<br>$A - F(I_{t-s}) - g(D_t)$ |

Given our assumption on the functions *F, G, f* and *g*, after some shot playing H will become the dominant strategy for the government, i.e., from that shot on the government starts playing the original version of the game as shown in Table 1. We assume that this happens in shot *T*.

Unless we assume that the oppositions are able to learn about the incompleteness of information after some shot, the lag irresponsibles that, although they asked the lockdown for *T* shots, the oppositions will suddenly start asking to end the quarantine for the following s shots (i.e., will continue saying the government is mistaken), wrongly computing the strategy that the government will play. Therefore, only at time *T + s* the oppositions will start supporting the government lockdown. In a more realistic fashion, the oppositions should start playing a Bayesian game, starting to assign subjective probability *p* and 1 − *p* to the government playing the game in Table 1 and 3 respectively. In this case, they could start playing E before shot *t* + *s*.

Formally, it must happen that:

(*i*) *D* − *f*(*I_t_*) − *g*(*D_t_*) < *C* − *F*(*I_t_*) − *G*(*D_t_*) for *t* < *T*, and vice-versa for *t* ≥*T*.

(*ii*) *C* − *f*(*I_t_*) − *g*(*D_t_*) < *A* − *F*(*I_t_*) − *G*(*D_t_*) for *t* < *T + k* and vice-versa for *t* ≥*T + k*.

Where *k* ≥ 0 reflects the fact that *A − C* ≥ *C − D*, i.e., the government might be slightly faster to switch preferences than the oppositions and the lobby.

In words, under incomplete information the government and the oppositions end up in the sub optimal Nash equilibrium (M,N) for the first *T* shots, then they start a transition towards the Pareto superior Nash equilibrium (H,E), staying in the sub optimal unstable configuration (H,N) for a number of shots between 1 and s according to the degree of learning of the oppositions, if any. As for the lobby, it will end up in a suboptimal configuration for *s + k periods*, while from periods 1 to *T* and *T + s + k* onwards it will be in its first best. This sub-optimality might last far more than in the model without information delays On the contrary, in case of complete information the government and the oppositions would stay in the sub optimal equilibrium up to a shot *τ* ≤ *T*, since at some point the Pareto superior equilibrium (H,E) might outweigh the risk-dominant suboptimal equilibrium (M,N) and from the following shot on there would be immediate convergence towards the payoff-dominant (H,E) without any costly convergence path. Moreover, the lobby would stay in its third best only from shot *τ +* 1 to shot *T + k*, where it should be reasonable to assume *T − τ −* 1 < *s*.

Since the exponential nature of our function *F* should set *T* and especially *k* to very low values, while the lag *s* is supposed to be up to two weeks, under complete information the society as a whole would set on the optimal path for far more shots than what happens under incomplete information. In some sense, the lobby is in its first best without knowing it.

### A2. Epidemiological model

The epidemiological model (flowchart in Figure A1) is as follows: (i) susceptible individuals (S) who acquire infection enter a latent phase where they are infectious but not infective yet; (ii) latency is represented through a sequence of *n* compartments characterised by the same exponentially distributed duration (*σ*), so that the overall distribution is of the latent phase is Gamma (*n, σ*); (iii) individuals leaving the last latency compartment become fully infective and can follow two distinct profiles, the asymptomatic (including pauci-symptomatic and non-specific symptoms) and the symptomatic profile, in proportions *f_A_ = f* and *f_S_ =* l *− f*. Asymptomatic infective will transmit the infection at a rate *β_A_*, recover at rate *γ_A_*, and are tested at a (possibly low) rate *τ_A_*. Upon testing, they are assumed to be isolated where they transmit at a lower rate *β_AC_ < β_AC_*, recover at rate *γ_AC_*, can be hospitalised at a (possibly low) rate *ν_AC_*. Symptomatic infective will initially enter a low/equivocal symptoms phase where they transmit infection at a rate *β_X_*, recover at rate *γ_X_*, and are tested at a rate *τ_X_*, upon which they enter the compartment of confirmed cases. Confirmed cases transmit at a lower rate *β_C_* (mirroring some degrees of isolation), recover at rate *γ_C_*, can be hospitalised at rate *ν_C_* (≫ *ν_AC_*), and die at a rate *μ_C_*. Hospitalised individuals (H) can either recover (at a rate *γ_H_*) or die (at rate *μ_H_*),d are assumed – just for sake of simplicity - to contribute negligibly to transmission.

**Figure A1.**
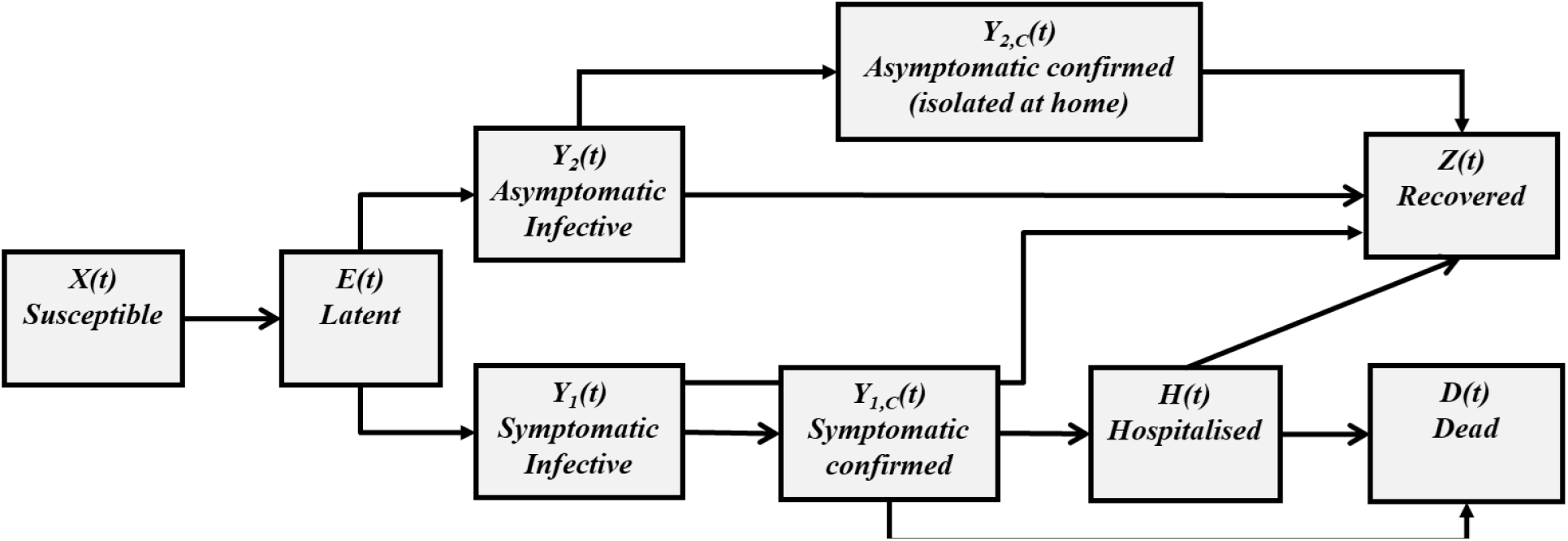
Flowchart of the adopted epidemiological model.

#### A.1. Time to lockdown (and unlocking) in some European countries

|  | Lockdown date | Epidemic size at lockdown (SOURCE: WHO) |  |  |  |
| --- | --- | --- | --- | --- | --- |
|  |  | Cumulated confirmed cases | New cases | Cumulated deaths | New deaths |
| Belgium | March 18<sup>th</sup> | 1486 | 401 | 14 | 9 |
| Denmark | March 18<sup>th</sup> | 977 | 79 | 4 | 3 |
| Italy | March 11-22 | 9172 | 1797 | 463 | 97 |
| France | March 17<sup>th</sup> | 7652 | 1079 | 175 | 27 |
| Germany | March 22<sup>nd</sup> | 21463 | 3140 | 67 | 22 |
| Norway | March 24<sup>th</sup> | 2371 | 239 | 8 | 1 |
| Spain | March 16<sup>th</sup> | 7753 | 2000 | 288 | 152 |
| UK | March 24<sup>th</sup> | 6654 | 967 | 335 | 54 |

## Footnotes

1 However, note that in a repeated game setting forward induction may lead the lobby to encourage the harsh policy if its support is critical in changing the policy implemented by the government in the next shot. This consideration is only one of the many cases demonstrating that the lack of political responsibility does not imply that the lobby does not play any role in influencing the decision of the other players.

